# Anterolateral entorhinal cortex thickness as a new biomarker for early detection of Alzheimer’s disease

**DOI:** 10.1101/19011825

**Authors:** Andrew Holbrook, Nicholas Tustison, Freddie Marquez, Jared Roberts, Michael A. Yassa, Daniel Gillen, for the Alzheimer’s Disease Neuroimaging Initiative

**Affiliations:** Department of Statistics, University of California, Irvine, CA, USA; Department of Radiology & Medical Imaging, University of Virginia, Charlottesville, VA, USA; Department of Neurobiology and Behavior and Center for the Neurobiology of Learning and Memory, University of California, Irvine, Irvine, CA, USA

**Keywords:** ADNI-1, Alzheimer’s disease, Anterolateral entorhinal cortex, Biomarker, Brain imaging, Clinical dementia rating, memory, Cortical thickness, CSF amyloid, Linear mixed-effects models, Mild cognitive impairment, Mini-mental state exam, Posteromedial entorhinal cortex, ROC

## Abstract

**Introduction:** Loss of entorhinal cortex (EC) layer II neurons represents the earliest AD lesion in the brain. Research suggests differing functional roles between two EC subregions, the anterolateral EC (aLEC) and the posteromedial EC (pMEC).

**Methods:** We use joint label fusion to obtain aLEC and pMEC cortical thickness measurements from serial MRI scans of 775 ADNI-1 participants (219 healthy; 380 MCI; 176 AD) and use linear mixed-effects models to analyze longitudinal associations between cortical thickness, disease status and cognitive measures.

**Results:** Group status is reliability predicted by aLEC thickness, which also exhibits greater associations with cognitive outcomes than does pMEC thickness. Change in aLEC thickness is also associated with CSF amyloid and tau levels.

**Discussion:** Thinning of aLEC is a sensitive structural biomarker that changes over short durations in the course of AD and tracks disease severity – it is a strong candidate biomarker for detection of early AD.

## Introduction

Layer II of the entorhinal cortex (EC) is one of the earliest sites for the accumulation of tangle pathology and neurodegeneration in the course of Alzheimer’s disease (AD) ^1–3^. Quantitative studies of neuron numbers in autopsy brains characterized for AD pathology have shown that a substantial reduction in EC is observed by the time of dementia diagnosis and further progressive loss of EC neurons occurs over the course of the disease ^4–6^. Little or no neuron loss occurs within EC in healthy aged brains without AD pathology suggesting that EC neurodegeneration is specific to disease ^4^.

Histopathological data indicate that the transentorhinal region, which consists of the anterolateral EC (aLEC) and perirhinal cortex, is vulnerable in the early stages of AD (Braak Stages I and II [2]). Recent evidence has elucidated a functional subdivision in the EC whereby the lateral and medial portions are involved in different aspects of information processing ^7^ and are differentially connected with the perirhinal and parahippocampal cortices ^8^. Other work has shown that the aLEC (which maps onto the lateral entorhinal cortex in rodents) is selectively vulnerable to age-related alterations in processing ^9^ as well as structural changes associated with age-related cognitive decline ^10^ in contrast to the posteromedial portion (pMEC). While volume reductions in the EC independently predict the likelihood of conversion from healthy aging to amnestic mild cognitive impairment (MCI) and from MCI to AD ^11–13^, preceding and predicting hippocampal volume reduction ^14^, it is unclear whether these volumetric changes are primarily driven by the aLEC or the pMEC.

Given the need for improved diagnostic biomarkers that are capable of detecting the earliest signs of neurodegeneration and the wealth of evidence pointing to the EC as an early site of structural decline, we seek to determine if we can identify different trajectories of structural thinning in the aLEC and pMEC in healthy, MCI and AD individuals.

The Alzheimer’s Disease Neuroimaging Initiative (ADNI ^15^) began in 2003 with the goal of developing imaging, genetic and pathological biomarkers for early detection and longitudinal progression in AD. This multisite imaging endeavor provides investigators with open access to serial MRI scans from nondemented individuals as well as MCI and AD patients, in conjunction with other biomarker data such as CSF amyloid and tau pathological markers. Measurements of cortical thickness (CT) have recently emerged as potential candidates for biomarkers due to their superior sensitivity to layer-specific cortical atrophy compared to volumetric approaches and the availability of automated methods for estimation ^16^. In the ADNI sample, EC CT was the most powerful measure of structural change both in MCI and AD brains ^17^. EC thinning also preceded and predicted hippocampal atrophy ^18^ and predicted conversion to AD with the greatest accuracy ^19^.

For EC thinning to be a reliable and robust measurement that accurately reflects neurodegeneration and supports longitudinal tracking of disease progression, several common methodological limitations need to be addressed ^20^. These issues include registration bias and inverse consistency, bias due to asymmetric interpolation favoring the baseline scan in longitudinal pipelines ^21^ and susceptibility to errors in segmentation or overestimation of gray matter thickness without specified anatomical constraints ^22^.

Here, we apply a novel pipeline that we recently developed for longitudinal registration-based CT to quantify aLEC and pMEC thinning that directly addresses these pitfalls and extend prior findings that EC thickness reliably differentiates normal controls from MCI patients and MCI patients from AD patients in the ADNI sample. Using linear mixed-effects (LME) models, we quantify cross-sectional and longitudinal associations between aLEC and pMEC thickness and two cognitive outcomes, the Clinical Dementia Rating – Memory box score (CDRM) and the Mini-Mental State Exam (MMSE), while controlling for possible confounding variables including age, sex, total brain volume and *APOE* ε4 genotype. We supplement this analysis of cognitive outcomes by using further LME models to establish diagnostic cohort specific trajectories in aLEC and pMEC CT through time and receiver operating characteristic (ROC) curves to ascertain predictive value of aLEC and pMEC CT for diagnostic outcomes. In a secondary analysis, we use an LME model to follow trajectories in aLEC and pMEC CT through time for two sub-cohorts with differing CSF amyloid profiles.

## Materials and Methods

### Raw imaging data and preprocessing

All T1-weighted MPRAGE MRI scans used in this study were drawn from the publicly available Alzheimer’s Disease Neuroimaging Initiative (ADNI). Exact parameters for the sequences acquired are available on http://adni.loni.usc.edu. Due to limited contrast between EC regions and surrounding areas in T1-weighted MRI, we employ the multi-atlas joint label fusion methodology ^23^ for EC parcellation and subsequent thickness estimation based on combined T1- and T2-weighted image information from a set of gold-standard atlases (see below), permitting a more robust weighted consensus approach than single-template and/or T1-weighted-only alternatives.

### Atlas data

We use a set of 17 atlases for multi-atlas joint label fusion comprising T1/T2-weighted image pairs and corresponding segmentation labels for the following left/right regions (aLEC, pMEC, perirhinal cortex, parahippocampal cortex, DG/CA3, CA1, and subiculum). Manual atlas labeling uses the T2-weighted image for each atlas set and a well-established and validated protocol ^9^. Atlas labels for a single subject are shown in **Supplementary Figure S1** superimposed on the corresponding T2-weighted image. The scans used to compose the atlases were collected on a Philips 3T scanner at the University of California, Irvine. T1-weighted MPRAGE scans were acquired in the sagittal orientation with an isotropic image resolution of 0.75 × 0.75 × 0.75 mm^3^. Image acquisition for the T2-weighted protocol was angled perpendicular to the long axis of the hippocampus consistent with previous work ^24^. T2-weighted image resolution is 0.47 × 0.47 × 2.0 mm^3^. The optimal rigid transformation between each individual atlas’ T1- and T2-weighted images was determined using the Advanced Normalization Tools (ANTs) software package ^25,26^.

### Population-specific templates

To facilitate aLEC/pMEC thickness estimation for the ADNI cohort described below, two population-specific, optimal shape/intensity templates were generated. The first T1-weighted template was constructed from 52 cognitively normal ADNI-1 subjects for a separate ADNI-based investigation ^27^, and we opted to use it in this study since it provides an intermediate registration space for transforming the labels of the 17 atlases. The second T1-weighted template, the “UCI” template, was generated from the 17 T1-weighted atlas images discussed above^28^. Representative slices for both templates are shown in **Supplementary Figure S2**. ANTs-based Symmetric normalization (SyN) was used to determine optimal diffeomorphic transformation between the two T1-weighted templates. This permits the two T1-weighted templates to act as an intermediate geometric space for the “pseudo-geodesic” mapping ^29^ between a set of atlas labels and the individual T1-weighted time point.

### Individual time point processing

Processing was conducted using the recently developed ANTs longitudinal structural processing pipeline ^27^ which is an extension of the previously reported cross-sectional framework ^30^. Briefly, the T1-weighted images constituting the set of subject’s longitudinal data were used to create a single-subject template (SST) as an unbiased space for processing longitudinal time points of individual subjects ^21^. The SST was then processed through the cross-sectional pipeline using the ADNI-1 template mentioned earlier. This processing produced the SST auxiliary images (i.e., *n*-tissue segmentation priors and brain extraction mask prior) used for individual time point brain extraction and tissue segmentation into CSF, cortical gray matter, white matter, deep gray matter, brain stem and cerebellum. Output of this processing stream includes the transforms between the individual time point and the SST and the transforms between the SST and the ADNI-1 template. In this way, concatenation of transforms can be used to map each of the 17 atlas label sets to each individual time point through a set of intermediary spaces which constitutes the “pseudo-geodesic” transform. This strategy has the benefit of reducing diffeomorphic distances between registration image pairs, reducing computational costs in terms of the sheer number of registrations, and taking advantage of the longitudinal nature of the data. This pseudo-geodesic mapping strategy is illustrated in **Supplementary Figure S3**.

### Multi-atlas joint label fusion

After mapping the set of 17 atlas label sets to each individual time point, the multi-atlas joint label fusion^23^ approach is applied. This technique weights the contribution of each atlas while minimizing informational redundancy between the atlases. To estimate CT for each EC region, we base our strategy on the MindBoggle approach^31^ but, instead of employing a mesh-based surface area calculation, we opt for the more accurate Crofton’s formula ^32^, which estimates the surface area directly.

### Statistical analyses

Our primary interest is the linear association between cognitive performance (CDRM and MMSE), diagnostic status (healthy, MCI and AD) and cortical thickness (CT) in the aLEC and pMEC. We seek to discern whether declining cognitive performance tracks with deterioration of CT within the two subregions. We also ask whether clinical diagnostic groups are separable when viewed through subregion CTs and their trajectories through time.

Linear mixed-effects (LME) ^33^ modeling allows us to leverage the longitudinal nature of the ADNI repeated-measures design insofar as a correctly specified LME model adjusts for within-subject correlation structure through time. As an extension of the multiple linear regression framework, LME modeling also supports adjustment for possible confounding variables as well as inclusion of precision variables. For the primary analysis, we use three LME models in total, each of which features subject-specific random intercepts and slopes through time. We decide on the inclusion of random components using the modified likelihood ratio test^34^.

With the first two models we wish to understand cognitive performance as a linear function of CT and its change through time. Both of these models regress either CDRM or MMSE over aLEC or pMEC CTs (and functions thereof) independently. We fit each model once for aLEC thickness as predictor of interest and once for pMEC thickness as predictor of interest since simultaneous inclusion of both measures results in multicollinearity on account of correlations between subregional CT. The first model evaluates cognitive score as a function of baseline thickness and the interaction between baseline thickness and months since baseline. The second model evaluates cognitive score as a function of baseline thickness and loss of thickness through time. We stratify the first two models by diagnostic cohort on account of the possibility of diagnosis based non-linearities in associations through time. Stratification decreases statistical power but increases model robustness.

Another primary question is whether population CT averages and their trajectories through time can be separated as a function of healthy, MCI and AD statuses. A third LME model (Model 3) independently regresses aLEC or pMEC CTs over diagnostic status and its interaction with months from baseline. We supplement Model 3’s inferential analysis with a predictive analysis using ROC curves ^35^ and area under these curves (AUC) to demonstrate prediction of diagnostic statuses using aLEC or pMEC thicknesses alone.

Given positive results, we motivate future research by asking the secondary question whether differential associations between CSF amyloid levels and aLEC/pMEC CTs provides explanatory power for primary analysis results. Based on prior work ^36–39^, Model 4 considers the ratio between p-tau and Aß binarized at the threshold 0.1 as predictor for CT in aLEC and pMEC subregions. All models are outlined in **Supplementary Table 1**. All modeling decisions were made prior to data access.

**Table 1.** Outcomes, predictors, and confounding variables. For each continuous variable, we show cohort means and standard deviations. For factors, we show the percentage of the cohort in each level. Baseline variables are shown with their natural scale, whereas change in these variables is shown using percentages to facilitate comparison across variables.

|  | Control (219) | MCI (380) | AD (176) |
| --- | --- | --- | --- |
| Baseline aLEC (mm) | 2.19 (0.14) | 2.11 (0.20) | 1.97 (0.19) |
| Loss aLEC (%/yr) | $6.7 \times 10^{-4}$ ($2.6 \times 10^{-2}$) | $1.1 \times 10^{-2}$ ($3.1 \times 10^{-2}$) | $1.3 \times 10^{-2}$ ($4.0 \times 10^{-2}$) |
| Baseline pMEC (mm) | 1.89 (0.13) | 1.85 (0.15) | 1.77 (0.16) |
| Loss pMEC (%/yr) | $1.4 \times 10^{-3}$ ($2.2 \times 10^{-2}$) | $5.2 \times 10^{-3}$ ($2.4 \times 10^{-2}$) | $6.9 \times 10^{-3}$ ($3.0 \times 10^{-2}$) |
| Baseline MMSE | 29.12 (0.97) | 27.06 (1.78) | 23.41 (2.04) |
| Loss MMSE (%/yr) | $4.0 \times 10^{-4}$ ($4.8 \times 10^{-2}$) | $2.7 \times 10^{-2}$ ($1.0 \times 10^{-1}$) | $1.0 \times 10^{-1}$ ($1.8 \times 10^{-1}$) |
| Baseline CDRM | 0.00 (0.15) | 0.57 (0.19) | 1.00 (0.32) |
| Gain CDRM (%/yr) | N/A | $1.7 \times 10^{-1}$ ($5.4 \times 10^{-1}$) | $2.5 \times 10^{-1}$ ($5.6 \times 10^{-1}$) |
| Brain volume (mm<sup>3</sup>) | $1.47 \times 10^6$ ($1.39 \times 10^5$) | $1.50 \times 10^6$ ($1.48 \times 10^5$) | $1.45 \times 10^6$ ($1.62 \times 10^5$) |
| Baseline age (yrs) | 75.97 (5.06) | 74.93 (7.14) | 75.01 (7.63) |
| APOE (% with (0, 1, 2) $\epsilon 4$ alleles) | (74, 24, 2) | (47, 42, 12) | (33, 48, 19) |
| Male (%) | 54 | 64 | 52 |

Continuous variables present as *mean (standard deviation)* aLEC: anterior lateral entorhinal cortex pMEC: posterior medial entorhinal cortex MMSE: mini-mental state exam CDRM: clinical dementia rating–memory APOE: apolipoprotein $\epsilon 4$

We use the R programming language ^40^ for all statistical analyses. We use the nlme package ^41^ for LME model fitting, the ggplot2 package ^42^ for visualization and the plotROC package for generating ROC curves ^43^. For exploratory analyses, we: present a data table with means, proportions and standard deviations of outcomes and model covariates stratified by diagnostic cohort; plot aLEC and pMEC thicknesses as a function of subject age, stratifying by sex; and use nearest neighbor missclassification as an index of homogeneity.

## Results

### Data distributions

We provide descriptive statistics for outcomes, predictors and other covariates in **Table 1** organized by diagnostic cohort. For each cohort, means and standard deviations appear for continuous variables and level-wise percent membership appears for factors.

For both baseline aLEC and baseline pMEC cortical thickness, the controls have the highest values, the AD cohort has the least, and the MCI cohort is in the middle. This trend holds for the longitudinal change in thickness. The AD cohort has the largest percent loss per year, and the MCI cohort has less percent loss per year. For both of these groups the %/yr loss is less for pMEC than it is for aLEC. MMSE and CDRM also follow the cohort-wise trends: baseline MMSE decreases from control cohort to AD cohort and baseline CDRM rises. For both MCI and AD cohorts, CDRM changes more through time than does MMSE.

**Figure 1** shows a scatterplot of unadjusted cortical thickness and age across sex and diagnostic cohort (healthy control and AD). **Figure 1a,c** shows aLEC thickness in males and females respectively, while **Figure 1b,d** shows pMEC thickness in males and females respectively. Visibly, there is greater overlap between healthy and AD cohort point clouds as a function of pMEC than as a function of aLEC. We quantify this overlap using the nearest neighbor misclassification rate as a homogeneity index. Regardless of sex, cohort clusters exhibit roughly 70% less homogeneity when viewed with aLEC thickness than with pMEC thickness.

### EC cortical thickness and cognitive performance

Models 1 and 2 regress cognitive performance over baseline and longitudinal CT. **Figure 2a** contains results from analyses based on Models 1 and 2. Green cells are nominally statistically significant at a 95% confidence level. Baseline CT and percent loss are standardized within cohort to facilitate cross-cohort comparisons and comparisons between the aLEC and the pMEC. In general, aLEC thickness is more predictive of outcome than is pMEC thickness. Across both outcomes (MMSE and CDRM), aLEC thickness has 8 significant associations with outcome, whereas pMEC only has 3 significant associations. In 9 of 12 of the comparisons shown in Table 2 effect sizes are larger for aLEC thickness.

**Figure 1.**
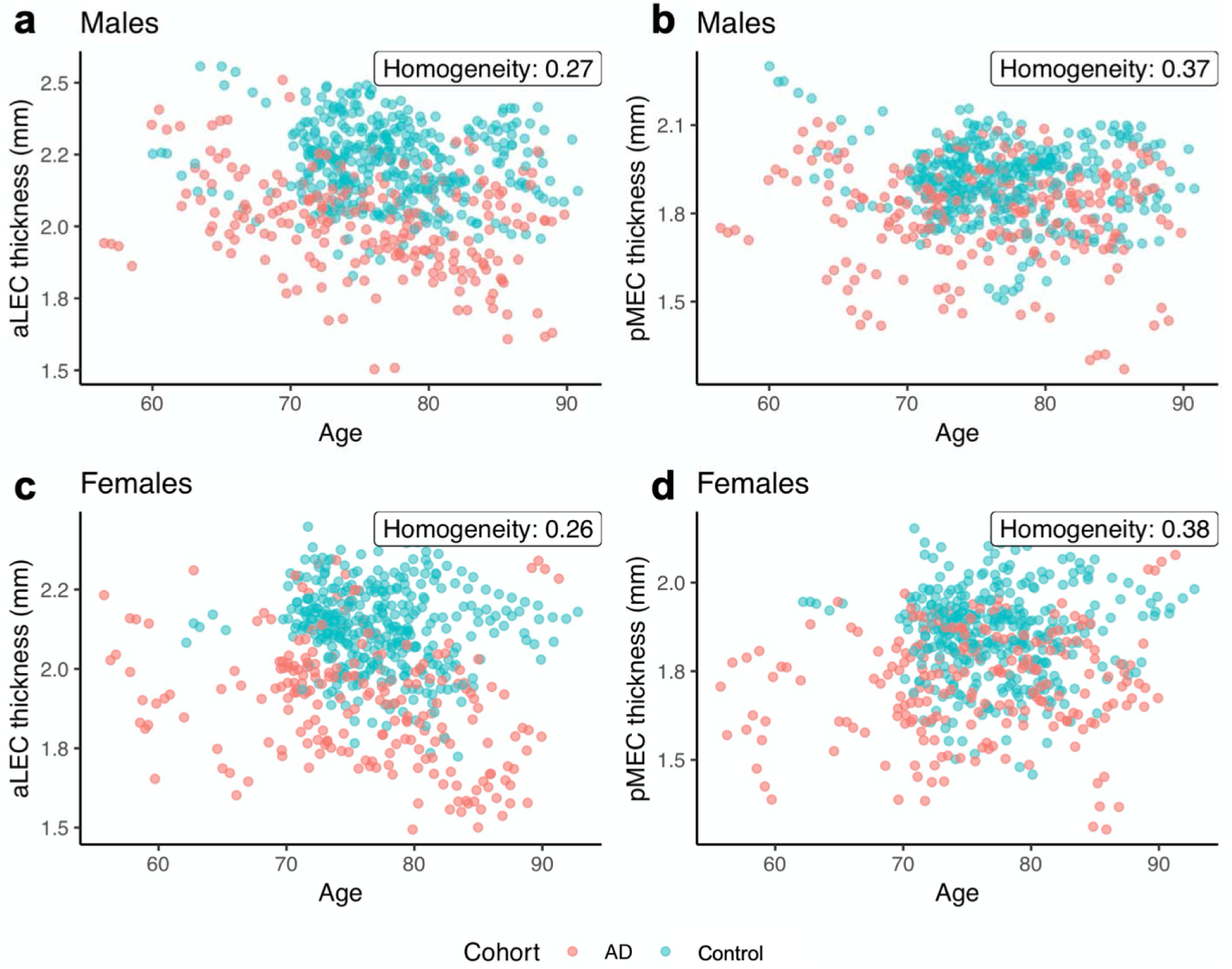
Scatterplots featuring anterolateral and posteromedial (aLEC and pMEC) cortical thickness (CT) and age stratified by sex and diagnostic cohort. aLEC thickness in males (a) and females (c) exhibits moderately less overlap between cohorts than does pMEC thickness in males (b) and females (d). We quantify overlap between healthy and AD cohorts using nearest neighbor misclassification rate as homogeneity index.

**Figure 2.**
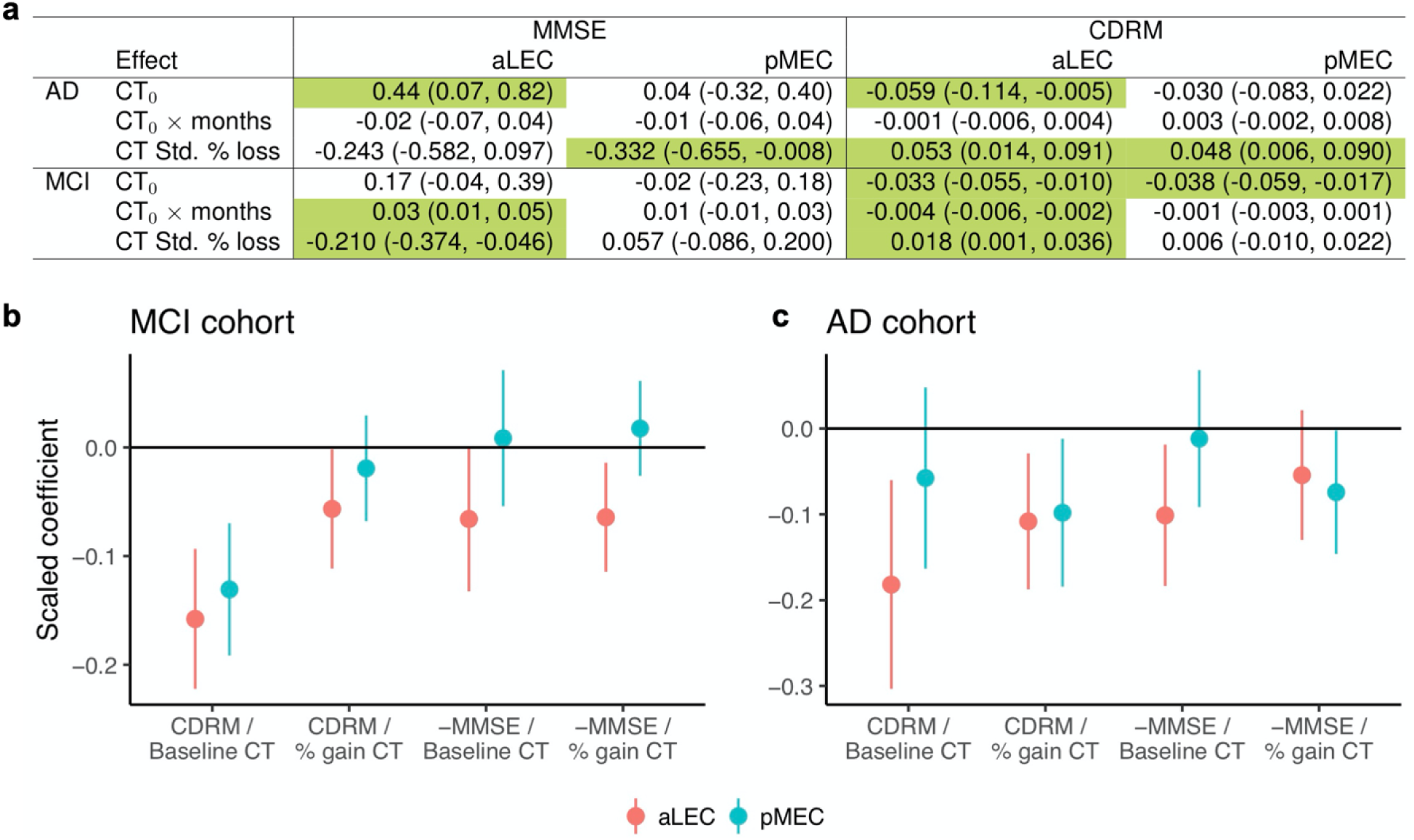
Estimated linear associations and nominal 95% confidence intervals between anterolateral and posteromedial entorhinal (aLEC and pMEC) cortical thicknesses (CT) and MMSE or CDRM. **(a)** for AD and MCI cohorts, the first row contains cross-sectional associations with baseline thickness (CT_0_) whereas the second and third lines contain longitudinal associations. Cells for which intervals do not contain zero are green. **(b-c)** Model 2’s adjusted linear associations between CDRM or MMSE and aLEC or pMEC baseline thicknesses and percent change in thickness from baseline. Baseline CT, percent gain CT, MMSE and CDRM are standardized. MMSE is negated since high performance is a higher score for MMSE but lower for CDRM. Associations are stronger for aLEC CT than for pMEC CT for both MCI **(b)** and AD **(c)**, exhibiting point estimates of greater scale as well as fewer confidence intervals overlapping zero.

**Figure 2** also illustrates Model 2 results, but, to facilitate comparisons across CDRM and MMSE and aLEC and pMEC thicknesses, axes are standardized. MCI cohort results are shown in **Figure 2b**, AD cohort results are shown in **Figure 2c**. We flipped the sign of MMSE so that lower scores reflect better testing performance for both cognitive measures. In general, regression coefficients reflecting the associations between CDRM or MMSE and aLEC (orange) thickness (and changes thereof) are more significantly non-zero than those of pMEC (blue) thickness. The scaled coefficients of aLEC are uniformly higher than pMEC except for the case of MMSE as a function of % loss CT for the AD cohort. For the MCI cohort, both lower baseline aLEC thickness and greater % loss aLEC CT predict worse CDRM and MMSE scores.

### EC cortical thickness and clinical diagnosis

Model 3 regresses CT over cohort membership and its interaction with time. In general, estimated effect sizes for aLEC as a function of cohort membership and time are twice those for pMEC. Nonetheless, all linear associations are nominally statistically significant at the 95% confidence level, i.e. none of the intervals contain zero.

The top row of **Figure 3** illustrates these results as a function of months from baseline. aLEC thickness is regressed over cohort membership and months in **Figure 3a**, pMEC thickness is regressed over the same in **Figure 3b**. The three cohorts exhibit greater separation at baseline when viewed through aLEC thickness than they exhibit when viewed through pMEC thickness. Estimated aLEC thickness 95% confidence bands maintain complete separation among cohorts throughout time, whereas estimated pMEC thickness 95% confidence bands do not.

**Figure 3.**
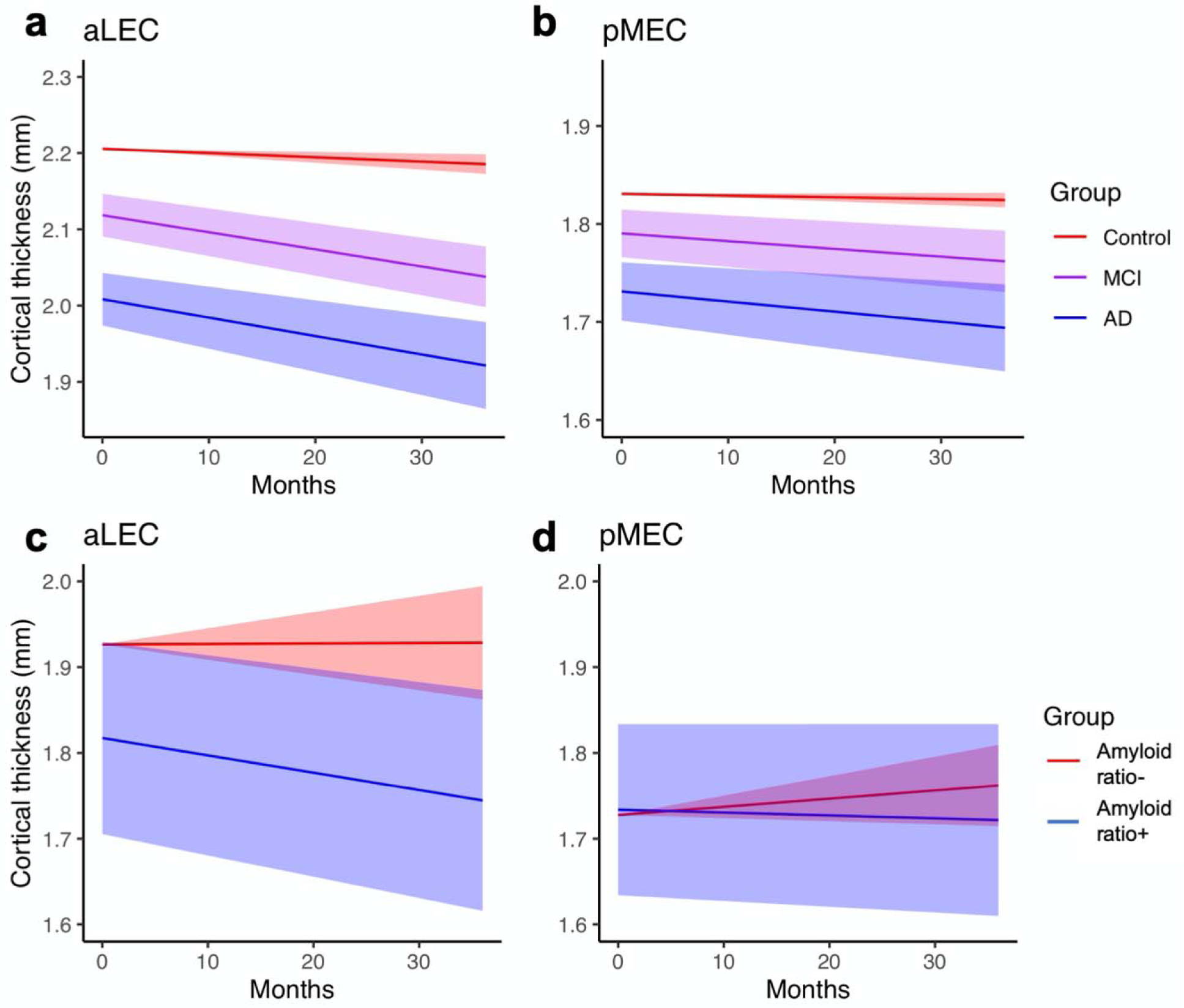
Subregion cortical thickness (CT) progressions through time as estimated using Model 3 along with 95% confidence bands. Model 3 accounts for individual variations as well as confounding variables. **(a-b)** when viewed through aLEC CT, the diagnostic cohorts exhibit statistically significant separation that persists through the entire time of measurement. Such separation is not apparent in pMEC CT. **(c-d)** secondary analysis on subset of ADNI-1 cohort comparing progressions for amyloid ratio-positive (p-tau/Aß >0.1) and ratio-negative cohorts shows qualitatively different behavior between aLEC and posteromedial entorhinal (pMEC) CT, suggesting a possible role for CSF amyloid ratio in influencing aLEC but not pMEC CT trajectory.

**Figure 4** supplements these inferential results with a predictive analysis using ROC curves to measure predictive content of aLEC and pMEC CTs with respect to MCI (**Figure 4a**) and AD (**Figure 4b**) status. The aLEC curves are consistently above the pMEC curves and yield higher AUCs, signifying greater predictive content at every threshold of the continuous CT values. Both aLEC and pMEC AUCs outperform those of subject age (MCI 0.47; AD 0.48) and total brain volume (MCI 0.47; AD 0.57).

### EC cortical thickness and CSF AD pathology

Given the stronger associations between aLEC CT and clinical outcomes than between pMEC thickness and the same, we ask whether a stronger link between aLEC thickness and CSF AD pathology levels exists than between pMEC thickness and the same. This secondary analysis provides a basis for future research into physiological mechanisms underlying aLEC CT and its clinical effects.

**Figure 4.**
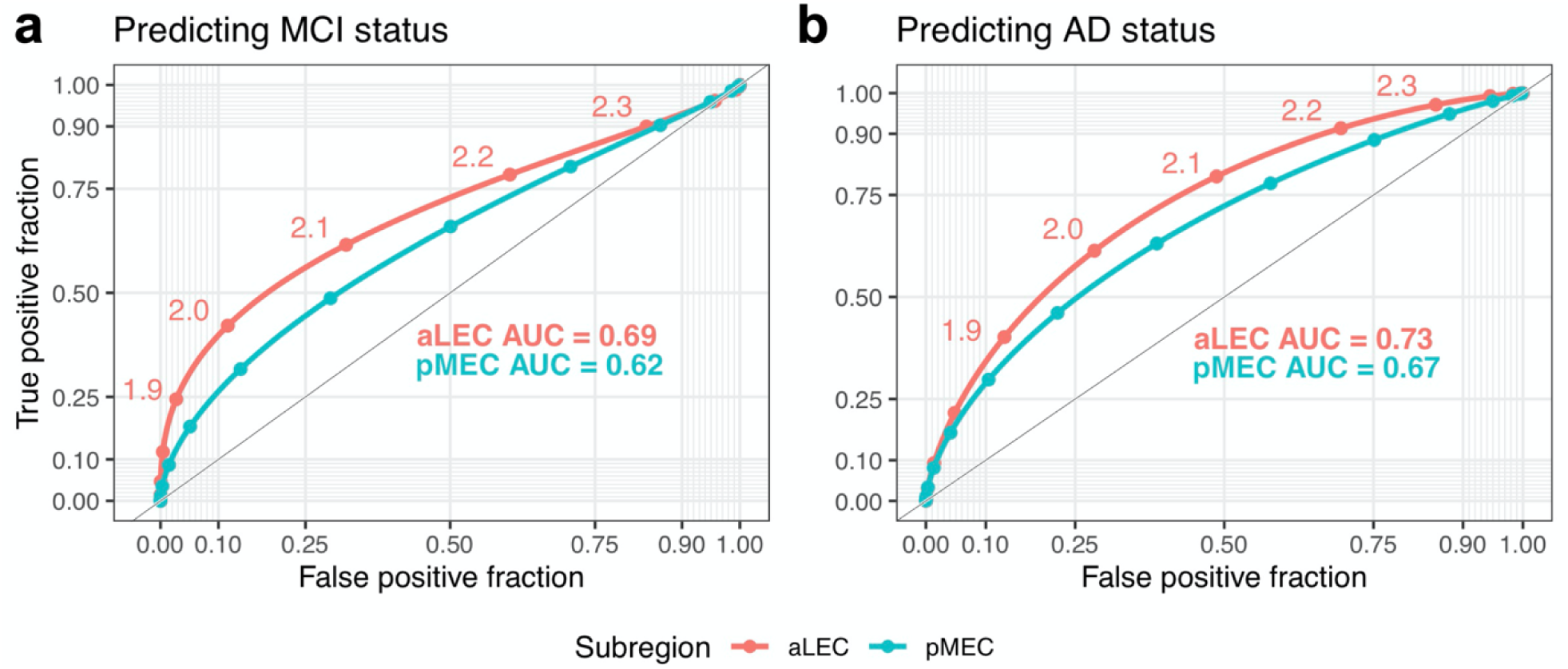
Receiver operating characteristic (ROC) curves for the prediction of MCI status and AD status using aLEC and pMEC CT. (a) In predicting MCI status, the aLEC curve dominates the respective pMEC curve and exhibits a larger area under the curve (AUC 0.69 vs. 0.62). (b) In predicting AD status, the aLEC curve also dominates the respective pMEC curve and exhibits a larger area under the curve (AUC 0.73 vs. 0.67). Both aLEC and pMEC AUCs outperform those of subject age (MCI 0.47; AD 0.48) and total brain volume (MCI 0.47; AD 0.57).

We look at the longitudinal progressions of aLEC and pMEC thicknesses as a function of the binary threshold given by the ratio of phosphorylated tau-181 (p-tau) to amyloid beta (Aß) being greater than 0.1 ^36–39^. These CSF data are available for a smaller 238 subject (70 healthy; 119 MCI; 49 AD) subset of the data used in the primary analyses. Due to dearth of repeated measures for CSF levels, we consider only the first CSF measurement for each individual and only include CT, CDRM and MMSE data collected during visits occurring after this CSF measurement with one-month grace period. Proportions of the ratio of p-tau to Aß that are greater than 0.1 are 0.9 for the healthy cohort, 0.97 for the MCI cohort and 1 for the AD cohort. We refer to these subjects as being “p-tau/Aß ratio-positive” or “amyloid ratio-positive”.

We model the linear associations between subregion CTs and ratio positivity and its interaction with time from baseline (as measured by time of CSF measurement). The bottom row of **Figure 3** presents the estimated linear cross-sectional (left) and longitudinal (right) associations along with 95% confidence intervals. Cross-sectionally, we estimate that the population of individuals with amyloid ratio positivity has 0.11 mm less aLEC CT than does the population of individuals who are amyloid ratio negative. For perspective, 0.11 mm is more than the difference between baseline aLEC thickness means of healthy control and MCI cohorts presented in **Table 1**.

Longitudinally, we estimate that the amyloid ratio-positive sample of individuals experiences an additional loss of 0.025 mm aLEC CT per year compared to the loss experienced by the amyloid ratio-negative sample. The additional loss in aLEC CT experienced by the amyloid ratio-positive sample requires 4 years before the difference between healthy and MCI cohorts is spanned. Due to the smaller sample size in this analysis, the results require further research and should be regarded as preliminary.

## Discussion

Given the wealth of research implicating the transentorhinal region^1–3^, selective vulnerability of the aLEC to age-related alterations in processing ^9^ and structural changes associated with age-related cognitive decline ^10^, we hypothesized that aLEC structure, specifically CT, might provide a suitable biomarker for early AD detection. We implemented a novel longitudinal CT pipeline on structural MRI data collected from the ADNI-1 cohort and compared this data with MMSE and CDRM performance, diagnostic cohort membership and CSF amyloid levels. Initial homogeneity analyses showed less overlap between healthy control and AD cohorts as a function of aLEC CT than for pMEC CT. We used LME models to analyze linear associations between these quantities through time while controlling for within-subject correlations and confounders such as age, sex, brain volume and *APOE* ε4 genotype.

Primary analyses showed statistically and practically significant negative associations between baseline aLEC thickness and progression of cognitive performance over time (Model 1). We also observed statistically and practically significant associations between change in aLEC thickness and cognitive performance through time (Model 2). Cross-sectional and longitudinal correlations between aLEC thickness and cognitive performance were present for both MCI and AD cohorts. We also tested whether trajectories of EC subregional CT through time differentiate by clinical diagnostic grouping (Model 3). aLEC thickness maintained complete separation between 95% confidence bands between healthy, MCI and AD cohorts while pMEC thickness did not.

Results indicate that the EC subregions could be differentially affected during early stages of AD. This is consistent with histopathological studies, which have reported that neurofibrillary tangles and neuropil threads show a distribution pattern that allow for staging ^3^. Initial stages show alterations confined to the transentorhinal region, which includes the aLEC. These results contribute to growing evidence that the aLEC is selectively vulnerable during early AD and also demonstrate that aLEC CT and changes in thickness over time are sensitive to cognitive changes and serve as a viable biomarker for prodromal AD.

In a secondary analysis, we analyzed the relationship between subregional CT and CSF measures of amyloid and tau pathology. Clinical symptoms of Alzheimer’s disease are preceded by a long preclinical phase in which pathological protein aggregation occurs in the brain ^6,44^. Additionally, Aβ plaques develop ∼15-20 years before onset of cognitive impairment and neurofibrillary tangles begin to accumulate at least 5 years before symptom onset ^44^. Previous studies have shown low CSF levels of Aβ strongly correlate with increased plaque load in the brain, and that high concentrations of CSF p□tau correlate with AD□specific neurofibrillary pathology ^45,46^. Furthermore, ptau_181_-Aβ_42_ ratio (ptau_181_/Aβ_42_) has been shown to be a strong predictor of conversion from cognitively normal to mild cognitive impairment over a 3∼4 year period ^36–38^.

We found statistically and practically significant linear associations between the binarized ratio p-tau/ Aβ >0.1 and aLEC CT and estimated that there are similar differences in aLEC CT levels comparing the p-tau/ Aβ ratio-positive sample to the ratio-negative sample as for the comparison between the MCI cohort and the healthy cohort. Furthermore, the p-tau/ Aβ ratio-positive sample exhibits a statistically and practically significant change in aLEC thickness over time, requiring an estimated 4 years to span the gap between healthy and MCI cohorts. This secondary analysis suggests the presence of AD-specific neuropathology may mediate thinning of the aLEC over time, but results require further investigation.

Overall, these results suggest that aLEC cortical thickness is a sensitive measure to cognitive decline as well as to AD pathological stage. Considering the growing interest in surrogate biomarkers that are sensitive and specific to AD especially during the early stages, we suggest that aLEC thinning may be an early marker that may be associated with cognitive decline especially in the memory domain and may serve as a mechanistic link between pathological load and cognitive outcomes. Additional research should focus on further understanding the function of aLEC and structural trajectories with aging and disease. For example, the human aLEC appears is involved in tasks ranging from visual object pattern separation ^7,9^ to intra-item configural processing ^47^ to temporal precision in real-world stimuli ^48^. Developing tasks that are specific and sensitive to aLEC (dys)function could serve as an early predictor of cognitive decline. In the future, these tasks can provide measures that can be used as neurobiologically-validated outcomes for clinical trials in preclinical AD.

## Data Availability

All data for this manuscript are available upon reasonable request.

## Acknowledgements

We acknowledge our sources of funding: T32 AG000096 (AH), NSF DGE-1321846 and B2D-1612490 (FM), as well NIA R01AG053555 and P50AG05146 (DG and MAY). We also acknowledge posthumously our co-author Jared Roberts who inspired and developed the initial stages of this project. Data collection and sharing for this project was funded by the Alzheimer’s Disease Neuroimaging Initiative (ADNI) (National Institutes of Health Grant U01 AG024904) and DOD ADNI (Department of Defense award number W81XWH-12-2-0012). ADNI is funded by the National Institute on Aging, the National Institute of Biomedical Imaging and Bioengineering, and through generous contributions from the following: AbbVie, Alzheimer’s Association; Alzheimer’s Drug Discovery Foundation; Araclon Biotech; BioClinica, Inc.; Biogen; Bristol-Myers Squibb Company; CereSpir, Inc.; Cogstate; Eisai Inc.; Elan Pharmaceuticals, Inc.; Eli Lilly and Company; EuroImmun; F. Hoffmann-La Roche Ltd and its affiliated company Genentech, Inc.; Fujirebio; GE Healthcare; IXICO Ltd.; Janssen Alzheimer Immunotherapy Research & Development, LLC.; Johnson & Johnson Pharmaceutical Research & Development LLC.; Lumosity; Lundbeck; Merck & Co., Inc.; Meso Scale Diagnostics, LLC.; NeuroRx Research; Neurotrack Technologies; Novartis Pharmaceuticals Corporation; Pfizer Inc.; Piramal Imaging; Servier; Takeda Pharmaceutical Company; and Transition Therapeutics. The Canadian Institutes of Health Research is providing funds to support ADNI clinical sites in Canada. Private sector contributions are facilitated by the Foundation for the National Institutes of Health (www.fnih.org). The grantee organization is the Northern California Institute for Research and Education, and the study is coordinated by the Alzheimer’s Therapeutic Research Institute at the University of Southern California. ADNI data are disseminated by the Laboratory for Neuro Imaging at the University of Southern California.

## Supplementary Materials

### Supplementary Methods

#### S.1. The ADNI dataset

Data used in the preparation of this article were obtained from the ADNI database (http://adni.loni.ucla.edu/). The ADNI was launched in 2003 by the National Institute on Aging (NIA), the National Institute of Biomedical Imaging and Bioengineering (NIBIB), the Food and Drug Administration (FDA), private pharmaceutical companies and non-profit organizations, as a $60 million, 5-year public-private partnership. The primary goal of ADNI (PI: Michael Weiner, UCSF) has been to test whether serial magnetic resonance imaging (MRI), positron emission tomography (PET), other biological markers, and clinical and neuropsychological assessment can be combined to measure the progression of MCI and early AD. Determination of sensitive and specific markers of very early AD progression is intended to aid researchers and clinicians to develop new treatments and monitor their effectiveness, as well as lessen the time and cost of clinical trials.

ADNI is the result of efforts of many co-investigators from a broad range of academic institutions and private corporations, and subjects have been recruited from over 50 sites across the U.S. and Canada. The initial goal of ADNI was to recruit 800 adults, ages 55 to 90, to participate in the research, approximately 200 cognitively normal older individuals to be followed for 3 years, 400 people with MCI to be followed for 3 years and 200 people with early AD to be followed for 2 years. A detailed description of the ADNI population, protocols and biomarkers is provided at http://adni.loni.ucla.edu/.

#### S.2. Subject selection

The ADNI general eligibility criteria are previously described ^49^. Normal controls (NC) have a CDR of 0. Subjects with MCI have a subjective memory complaint, objective memory loss measured by education-adjusted scores on Wechsler Memory Scale Logical Memory II, a CDR of 0.5, preserved activities of daily living, and absence of dementia. Subjects with AD have a CDR of 0.5 or 1.0 and meet NINDS criteria for probable AD. At the time of download, 300 individuals with MCI and 191 healthy controls with baseline, 6 months, and 12-month follow-up data were available for download and were used in the current study. We also randomly selected a sample of 49 AD patients with baseline scans for comparison and to define the parametric space of EC thickness. Demographics and baseline neuropsychological variables for all three groups are shown in **Table 1**. Neuropsychological test data were not available for 4 AD subjects.

#### S.3. MRI methods

Detailed methods of MRI acquisition are previously described ^50^. Only T1-weighted 3D MP-RAGE scans were used in this report (acquisition parameters: FOV = 240 × 240; matrix = 192 × 192; TR = 3000 ms; TI = 1000; flip angle = 8 degrees, slice thickness = 1.2 mm; sagittal orientation). All MPRAGE scans underwent quality control procedures, N3 bias correction and were scaled for gradient drift using phantom data.

#### S.4. Inferential model building and variable selection

To moderate inflation of type 1 error, we design our statistical models from first principles and prior to accessing the data. We select model responses and predictors of interest based on our neuroscientific questions of interest. After this, we decide upon the inclusion of additional covariates based on whether they might be confounders, i.e., variables that might influence both outcome and predictors of interest, or precision variables, i.e., variables that influence outcome alone. Inclusion of confounders decreases estimator bias and increases variance, leading to more conservative intervals. Inclusion of precision variables tightens confidence intervals, increasing certainty.

With these relationships in mind, we now discuss inclusion rationale. The first two models regress cognitive indices over CT and functions of CT and time from baseline. As such, we include *months* from baseline, clinical *diagnosis*, the *number of APOE* ⍰*4 alleles, sex, age* and *total brain volume* as potential confounding variables – months from baseline plausibly modulates CT and cognition scores; as a primary biomarker for genetic predisposition, APOE allele count certainly associates with cognition and might associate with CT; when combined with other subject descriptors, sex might modulate cognitive performance and certainly modulates CT; age certainly modulates both; and brain volume plausibly modulates both. For Model 3, similar logic applies by exchanging clinical diagnosis for cognition scores. A difference is that we take months from baseline to be a precision variable since CT changes with time but diagnosis remains constant for the cohort we consider.

Importantly, we adjust for all but one potential confounder by inclusion in the regression models as covariates, with the only exception being diagnostic status for Models 1 and 2. Since diagnostic differences plausibly modify the relationship between cortical thickness and MMSE/CDRM in complicated, nonlinear ways, we instead stratify the analysis by diagnosis, fitting the two models to the individual cohorts separately. Such stratification increases model robustness but decreases power, here expressed as wider, more conservative confidence intervals. Variables of all models appear in **Table S1**.

## Supplementary Results

### Statistical interpretations for Models 1 and 2

We provide statistical interpretations here as examples. Focusing on the upper left cell of Figure 2 in which the association between baseline aLEC thickness and MMSE is presented), the interpretation is that for every additional standard deviation in baseline aLEC thickness, there is an estimated gain of 0.44 (95% CI: 0.07, 0.82) MMSE expected for the AD subjects. For the result in which CDRM is modeled as a function of the interaction between baseline aLEC cortical thickness and months from baseline within MCI subjects, our interpretation is that for every additional standard deviation of baseline aLEC thickness and for each additional month from baseline, there is an estimated decrease of 0.004 (95% CI: 0.002, 0.006) in CDRM expected for MCI subjects. Finally, for the estimated linear association between standardized percent loss aLEC thickness and CDRM for the AD population, our interpretation is that for every additional standard deviation of percent loss from baseline, there is an estimated increase in CDRM of 0.053 (95% CI: 0.014, 0.091) expected for AD subjects.

### Statistical interpretations for Model 3

Considering aLEC thickness as a function of AD group membership and its interaction with time, it is estimated that when comparing the AD group to the healthy group, (1) the AD group has 0.20 mm (95% CI: 0.16, 0.23) lower aLEC thickness expected, all other covariates being held equal; and (2) the AD group has 0.02 mm (95% CI: 0.01, 0.04) lower aLEC thickness expected for each additional year from baseline, all other covariates being held equal. For the MCI group, estimated cross-sectional association with aLEC thickness is half that of the AD population (0.9 mm; 95% CI: 0.06, 0.12), but the estimated longitudinal association is roughly equal to that of the AD population (0.02 mm; 95% CI: 0.01, 0.02).

**Table S1.** Linear mixed-effects models and their variables.

| Model | Variable type | Variables |
| --- | --- | --- |
| I | Response | MMSE or CDRM |
|  | Predictor of interest | CT<sub>0</sub> and CT<sub>0</sub>×months for aLEC or pMEC |
|  | Potential confounders | months, diagnosis, APOE, sex, age, brain volume |
|  | Precision variable | — |
| II | Response | MMSE or CDRM |
|  | Predictor of interest | CT<sub>0</sub> and % loss CT for aLEC or pMEC |
|  | Potential confounders | months, diagnosis, APOE, sex, age, brain volume |
|  | Precision variable | — |
| III | Response | CT for aLEC or pMEC |
|  | Predictor of interest | diagnosis (control, MCI, AD) |
|  | Potential confounder | MMSE, APOE, sex, age, brain volume |
|  | Precision variable | months |
| IV | Response | CT for aLEC or pMEC |
|  | Predictor of interest | p-tau/Aβ > 0.1 |
|  | Potential confounder | MMSE, APOE, sex, age, brain volume |
|  | Precision variable | months |

CT: cortical thickness CT<sub>0</sub>: cortical thickness at baseline aLEC: anterior lateral entorhinal cortex pMEC: posterior medial entorhinal cortex MMSE: mini-mental state exam CDRM: clinical dementia rating–memory APOE: apolipoprotein ε4 p-tau: phosphorylated tau-181 Aβ: amyloid beta 42

**Figure S1.**
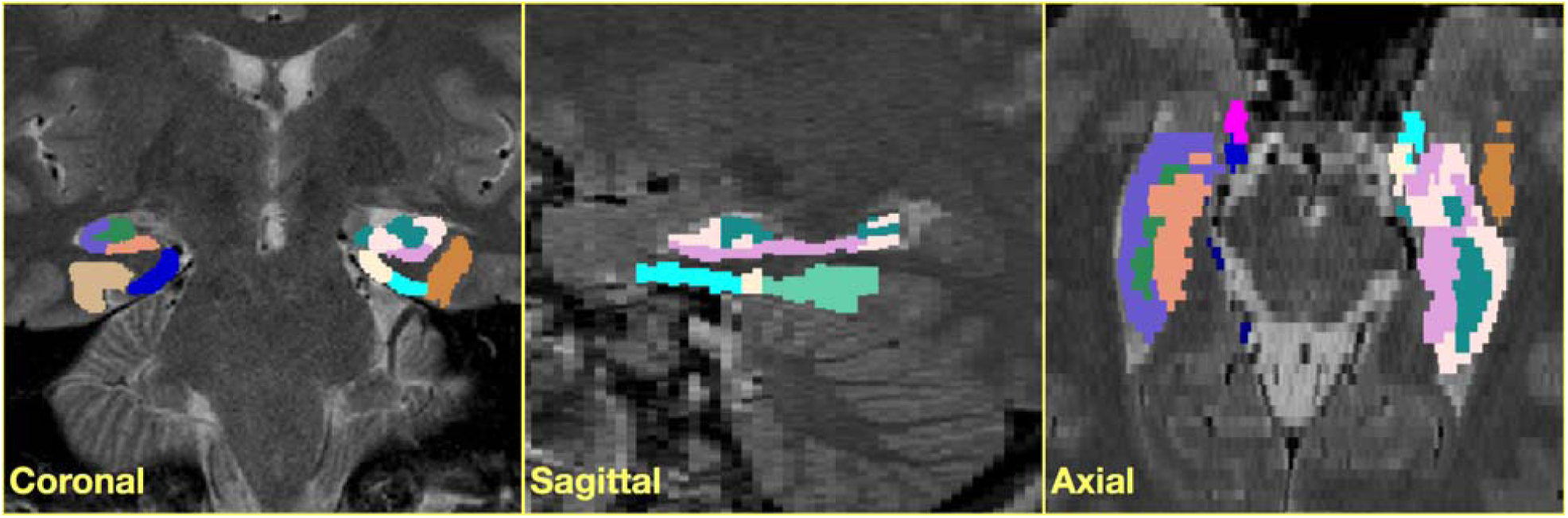
Atlas labels for Subject S1 partitioning the EC/hippocampal cortical complex. Each set of labels for the 17 subjects was manually placed in the space of the subject’s T2-weighted image using the procedure specified in the text.

**Figure S2.**
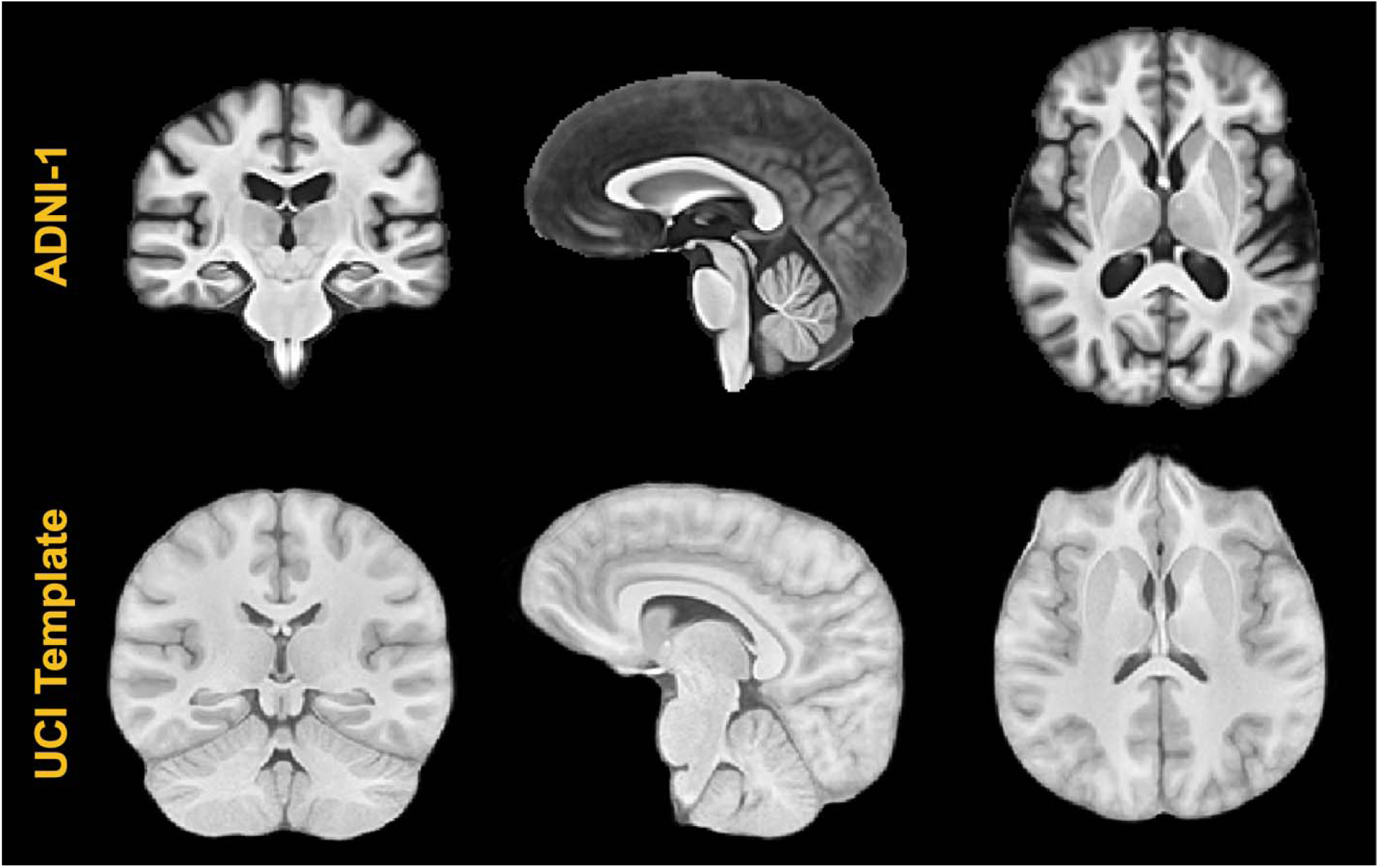
Representative views of the two population-specific templates created for this study. The ADNI-1 template was created from 52 cognitively normal subjects selected from the ADNI-1 template while the UCI template was created from the 17 T1-weighted images of the atlas set used for joint label fusion. These images constitute the intermediate spaces for the pseudo-geodesic transform between the EC labels and the T1-weighted images representing individual subject time points.

**Figure S3.**
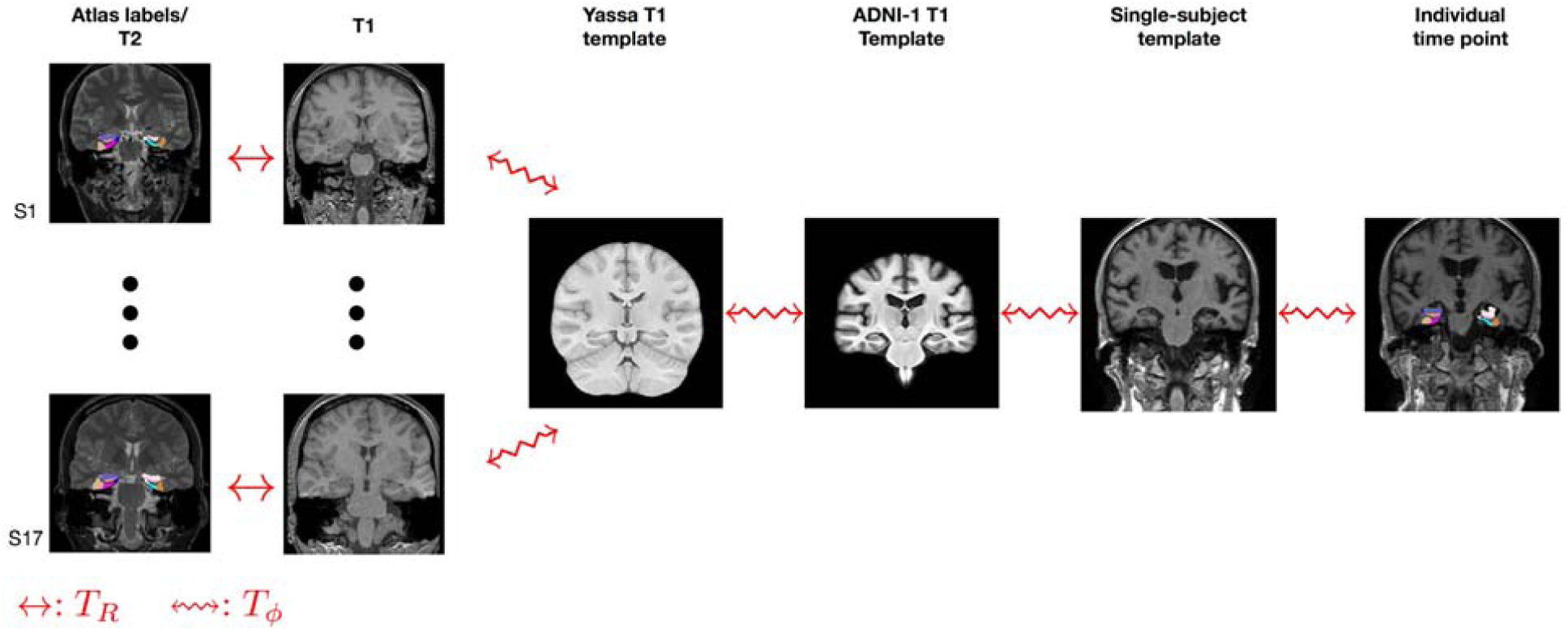
Illustration of the set of transforms used to map the set of 17 atlas labels to the T1 image of each individual time point. This pseudo-geodesic scheme minimizes the total number of pair-wise registrations for this study while taking advantage of the longitudinal aspect of the data. T_R_ and T_□_ denotes rigid and diffeomorphic transforms, respectively.

